# IMPACT OF PHYSICAL ACTIVITY ON POSTURAL STABILITY AND COORDINATION IN CHILDREN WITH POSTERIOR FOSSA TUMOR: RANDOMIZED CONTROL PHASE III TRIAL

**DOI:** 10.1101/2022.07.26.22277976

**Authors:** Mahmoud Usama, Faten Abdelaziem, Wafaa M. Rashed, Eslam Maher, Mohamed El Beltagy, Wael Zekri

## Abstract

**Background:** Posterior fossa tumor is a type of brain tumor that is located at the borders of both the brain stem and cerebellum. The cerebellum is the brain region in charge of balance and coordination. Pediatric patients diagnosed with posterior fossa tumor have been reported to fall frequently.

**Objectives:** The aim of this study is to investigate the effectiveness of balance and coordination training in these children.

**Methods:** This randomized control clinical trial (ClinicalTrials.gov Identifier: NCT04528316) was carried out between September 2020 and April 2021 at Children’s Cancer Hospital-57357. The inclusion criteria were - patients with posterior fossa tumor in maintenance phase and, age between 5-12 years. The exclusion criteria were - patients who had a genetic disorder or suffer from mental retardation, a chronic lung disease, sever cardiomyopathy, or a neuromuscular disease that doesn’t not related to tumor. The study participants were randomly assigned into three groups: Group I/Control group: they received Pilates core stability exercises program, Group II/Postural stability group: They received the same program plus HUMAC balance program, and Group III/Coordination group: They received the same program plus coordination exercises of BOT-2. The semi-parametric proportional odds model was used to compare follow up scores of the Postural stability group vs Control, and Coordination group vs Control, while adjusting for baseline values. All tests were two-sided, with alpha set to 0.05.

**Results:** Sixty children including 38 boys and 22 girls were enrolled in this study. In all three groups, postural stability and coordination improved significantly in terms of modified clinical test of sensory integration of balance, center of pressure, limits of stability, bilateral coordination, and upper-limb coordination.

**Conclusion:** The current study supports the value of adding postural stability and coordination training to the physiotherapy plan for children with posterior fossa tumor.

## 1. INTRODUCTION

Primary brain tumors account for nearly 20% of all malignant diseases in children and adolescents worldwide.^1^ Based on hospital-based cancer registry, brain tumors represent 18.12% of all malignant tumors at Children’s Cancer Hospital Egypt 57357 (CCHE 57357) and posterior fossa tumors (including the brainstem) represent 55.7% of brain tumors (data not published). These tumors rank second among all childhood cancers, second only to leukemia, and are the most common solid tumor in children.^2^

Post-tumor rehabilitation and overcoming the natural side effects of life-sustaining treatment are the main challenges facing neuro-oncology nowadays.^3^ Posterior fossa tumor arises within the cerebellum and spreads rapidly via cerebrospinal pathways forming tumors of various size alongside the ventricles.^4^ The treatment depends on three poles: surgery, chemo- and radiotherapy.^5^ The advances in these methods have improved survival rates, although the tumor itself and treatment options can cause undesirable side effects such as hearing loss, cognitive impairment and movement disorders.^6^ Posterior fossa tumor negatively affects sensorimotor function, balance control, vestibular reflexes and postural ability.^7^ However, the degree and type of motor disorders has not been fully investigated.

Postural balance is the ability to maintain the line of gravity related to the individual’s base of support, with minimal postural fluctuations, which requires good motor function and effective interaction between somatosensory, visual and vestibular systems.^8^ Balance requires multisystem feedback like intact cerebellar and cognitive functions for achieving proper balance, cognitive processing is essential in the process of postural control even in simple tasks like quiet standing as the motor movement needs expectation, attention, intention and experience, it is believed that the harder the task the more cognitive ability it would need.^9^

Poor posture control and imbalance are common defects after surgical resection of brain tumors. The posture and balance theory based on the neural control system shows that the musculoskeletal system and the nervous system work together to maintain normal postural control through individual interaction with tasks and environment.^10^

The inconsistency of available data makes the prescribed rehabilitation of improving postural balance a very difficult task. Several authors reported two cases with positive results after 8-week intervention, including neuromodulation devices as part of intensive physical therapy.^11^ Others have demonstrated the effectiveness of a 12-week intervention including stability of 20 cases exercise under the same conditions.^12^ However, a small sample size, as well as the lack of consensus on rehabilitation plans do not allow positive results to be explicitly transferred to other survivors after posterior fossa tumor.^13^

Coordination of many body parts to achieve smooth movements is generally agreed to be the specific role of cerebellar control. It is generally believed that this is due to the “fine tuning” of many motor pattern generators below the cerebellum in the spinal cord, brain stem, and motor cortex. There is an additional mechanism for coordination of body parts in posture and movement.^14^

Thus, to overcome previous limitations, the current study aimed to investigate the effectiveness of postural stability and coordination training in these Children.

## 2. METHODS

### 2.1. Trial participants

We conducted randomized control clinical trial at Children’s Cancer Hospital Foundation 57357, Cairo between September 2020 to April, 2021. The inclusion criteria include: 1-Children with posterior fossa tumors during follow up, 2-The age of study participants is between 5-12 years old, 3-time elapsed since the start of treatment is more than 4 months in order to be in maintenance phase, 4-they can understand verbal command, 5-children with no visual, auditory or perceptual disorders. The exclusion criteria include patients who had a genetic disorder or suffer from mental retardation, a chronic lung disease, sever cardiomyopathy, or a neuromuscular disease that doesn’t not related to tumor.

Ninety-two children with posterior fossa tumor participated in the study, but only sixty children treated for posterior fossa tumor met the eligibility criteria as illustrated in Figure 1. The study was approved by the institutional review board (IRB) at CCHE 57357 (CCHE-IRB:6/2020) (CHHE-IRB:10/2021). This clinical trial registered on ClinicalTrials.gov (ClinicalTrials.gov Identifier: NCT04528316). Informed consent was obtained from parents of all participants before starting procedures. All patients’ information was secured in REDCap Database (https://Pubmed.ncbi.nlm.gov/18929686) at CCHE-57357.

**FIGURE 1.**
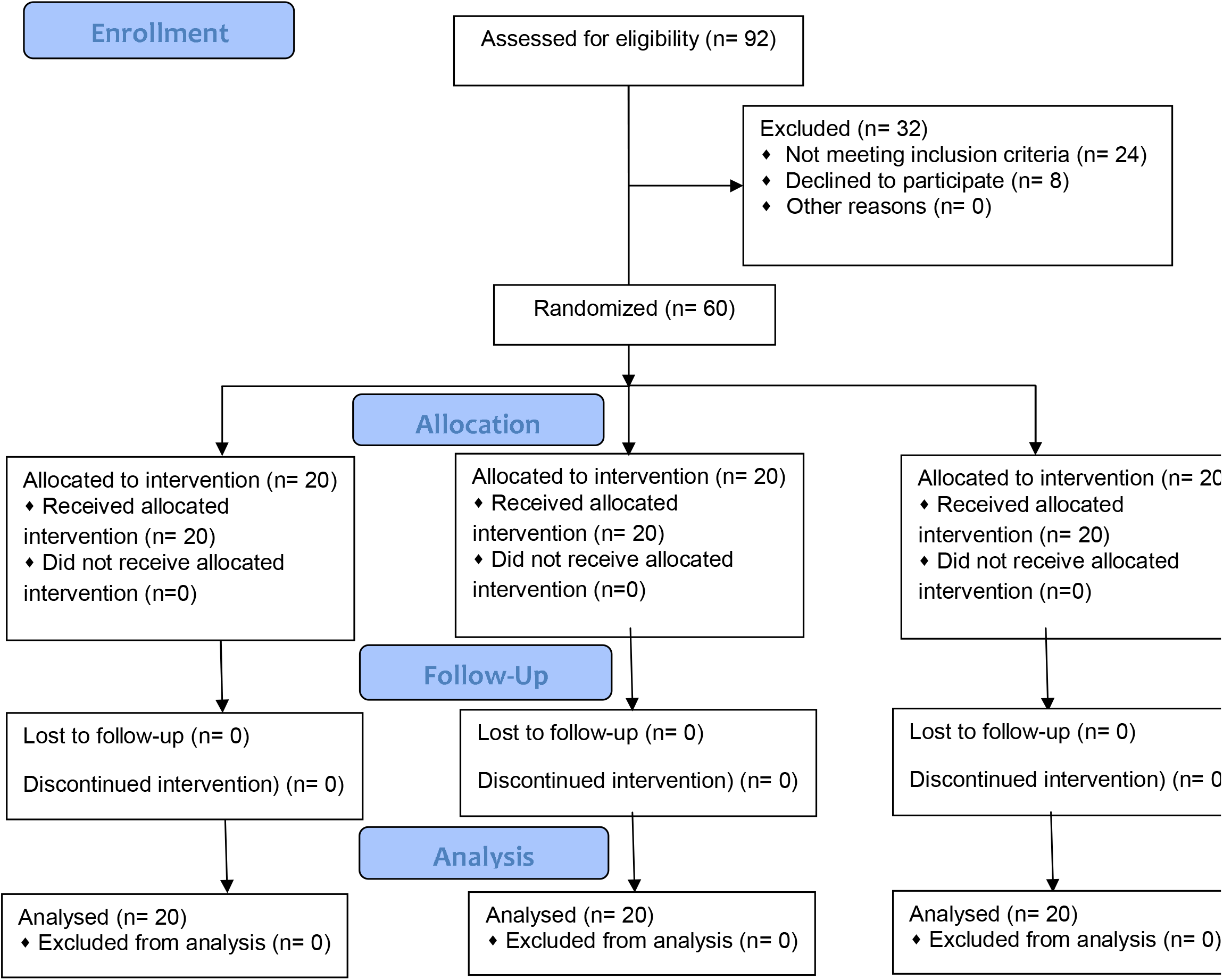
Flow diagram illustrating participants entered the study.

### 2.2. Procedures and measures

This randomized clinical trial includes three parallel groups:

#### Control group

This group received Pilates core stability exercises program for 60 minutes.^15^

#### Postural stability group

This group received HUMAC balance and tilt program for 30 minutes in addition to the control group’s selected program to examine the effect of using HUMAC balance and tilt system. The HUMAC software was designed to work with the mCTSIB (modified clinical test of sensory integration of balance) with eyes open and eyes closed, the centre of pressure (COP), and the limit of stability (LOS).^16^

#### Coordination group

This group was given coordination exercises from BOT-2 with repetitions for 30 minutes in addition to the control group’s selected program for 30 minutes to investigate the effect of using coordination exercises.^17^

### 2.3. Sample size

In an analysis of covariance analysis (ANCOVA), a total sample size of 60 is needed for the three arms combined, with equal allocation ratio, to power a minimum post-intervention score mean difference of 5, corresponding to a size of variation in the means of 2.36 for their standard deviation. The common standard deviation within a group is assumed to be 10, alongside a medium R-squared of 0.7 for the baseline mean (the covariate). This gives a beta error rate of 0.186 to detect differences among the means versus the alternative hypothesis of equal means using an F test at 0.05 alpha error rate.^18^

### 2.4. Randomization

Subjects were randomly assigned into three groups of equal number. Randomization was carried out at clinical trials unit-Research Department at CCHE 57357 using block-stratified randomization software (Rand.exe version 6).

### 2.5. Statistical analysis

The median (IQR) was used to summarize postural stability scores. The semi-parametric cumulative probability (proportional odds) model was used to compare follow up scores of the Balance group vs Control, and Coordination group vs Control, while adjusting for baseline values. All tests were two-sided, with alpha set to 0.05. Multiplicity adjustment was done using the Bonferroni correction method. R version 3.6.1 and IBM SPSS v20 were used.

## 3. RESULTS

### 3.1. Clinical characteristics of study participants

Demographic and clinical characteristics are summarized in Table 1.

**TABLE 1.** Demographic and clinical characteristics

### 3.2. Comparison between groups post treatment

There was significant difference between postural stability and control groups in postural stability and coordination scores. There was no significant difference between coordination and control groups in postural stability scores except for Eyes Open Firm Surface (EOSS), but there was significant difference between them in coordination scores.

#### 3.2.1. Postural stability scores

##### Modified clinical test of sensory integration of balance (mCTSIB)

Eyes Open Firm Surface (EOSS) in control group has a median score of 88 (IQR=86 – 88.5), in postural stability group has a median score of 90 (IQR=89 – 90) with adjusted [P-value< 0.0001] and in coordination group has a median score of 85.5 (IQR=85 – 89) with adjusted [P-value = 0.019]. Eyes Closed Firm Surface (ECSS) in control group has a median score of 80 (IQR=79.5 – 85), in postural stability group has a median score of 87.5 (IQR=85 – 88) with adjusted [P-value = 0.0002] and in coordination group has a median score of 79.5 (IQR=76.5 – 86.5) with adjusted [P-value = 0.92]. Eyes Open Foam Surface (EOFS) in control group has a median score of 68.5 (IQR=79 – 90), in postural stability group has a median score of 80 (IQR=77 – 85) with adjusted [P-value < 0.0001] and in coordination group has a median score of 72.5 (IQR=62.5 – 78) with adjusted [P-value = 0.43]. Eyes Closed Foam Surface (ECFS) in control group has a median score of 49 (IQR=35 – 70), in postural stability group has a median score of 71.5 (IQR=67.5 – 75) with adjusted [P-value < 0.0001] and in coordination group has a median score of 60 (IQR=50 – 68) with adjusted [P-value = 0.16].

##### Center of pressure (COP)

Center of pressure (COP) in control group has a median score of 85.5 (IQR=85– 89.5), in postural stability group has a median score of 93 (IQR=89 – 95) with adjusted [P-value < 0.0001] and in coordination group has a median score of 90 (IQR=88 – 90) with adjusted [P-value = 0.15].

##### Limits of stability (LOS)

Limits of stability (LOS) in control group has a median score of 36 (IQR=30– 40), in postural stability group has a median score of 51.5 (IQR=44 – 54) with adjusted [P-value< 0.0001] and in coordination group has a median score of 40 (IQR=30.5 – 44) with adjusted [P-value = 0.56].

#### 3.2.2. Coordination scores

##### Bilateral coordination

Bilateral coordination in control group has a median score of 18 (IQR=17.5– 19), in postural stability group has a median score of 20 (IQR=19 – 20) with adjusted [P-value = 0.0012] and in coordination group has a median score of 22 (IQR=21 – 22) with adjusted [P-value < 0.0001].

##### Upper limb coordination

Upper limb coordination in control group has a median score of 25 (IQR=22– 31.5), in postural stability group has a median score of 30 (IQR=29.5 – 32) with adjusted [P-value = 0.005] and in coordination group has a median score of 36 (IQR=35 – 37) with adjusted [P-value < 0.0001]. (Table 2, Fig. 2)

**TABLE 2.** Comparison between groups post treatment

**FIGURE 2.**
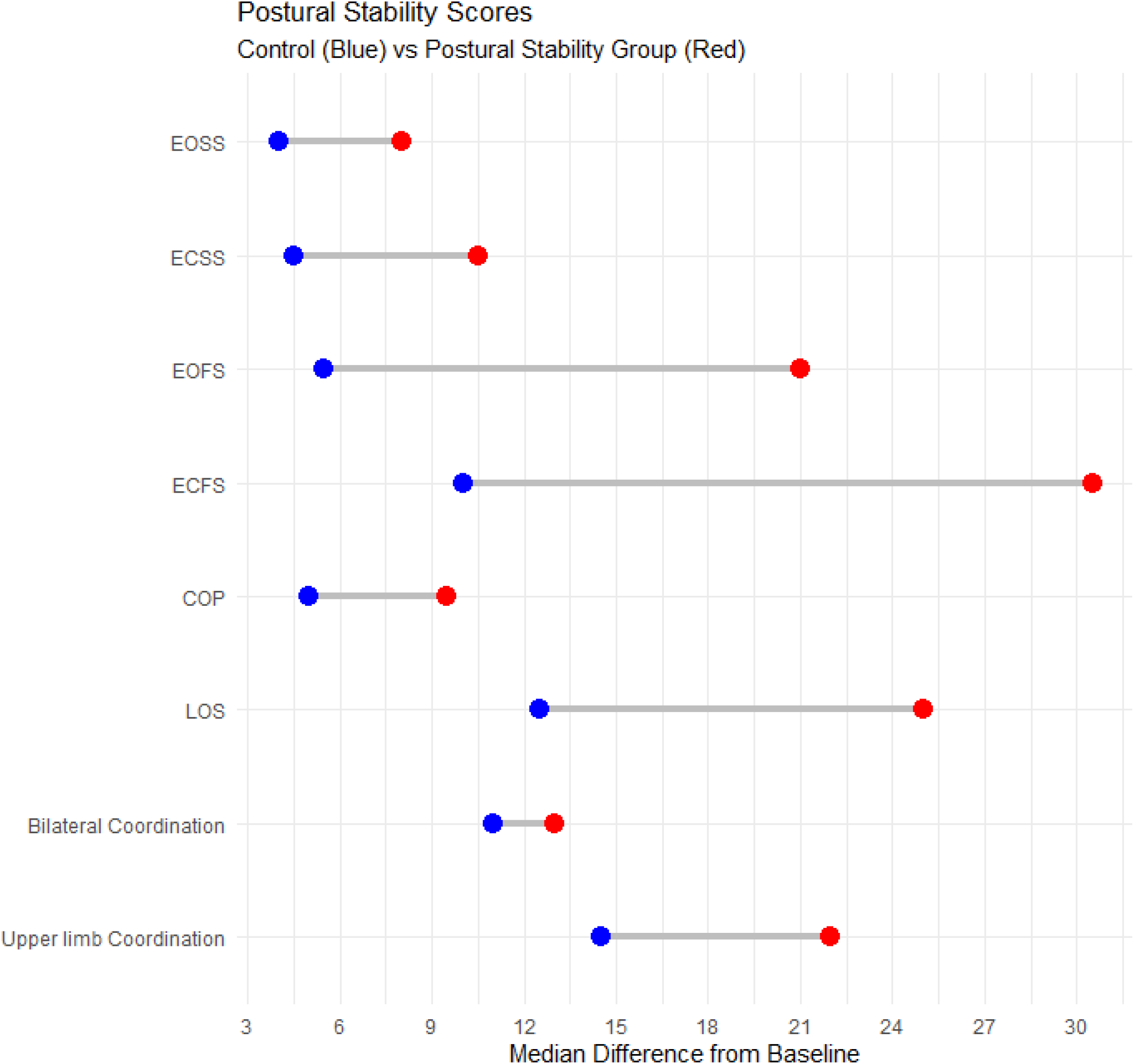
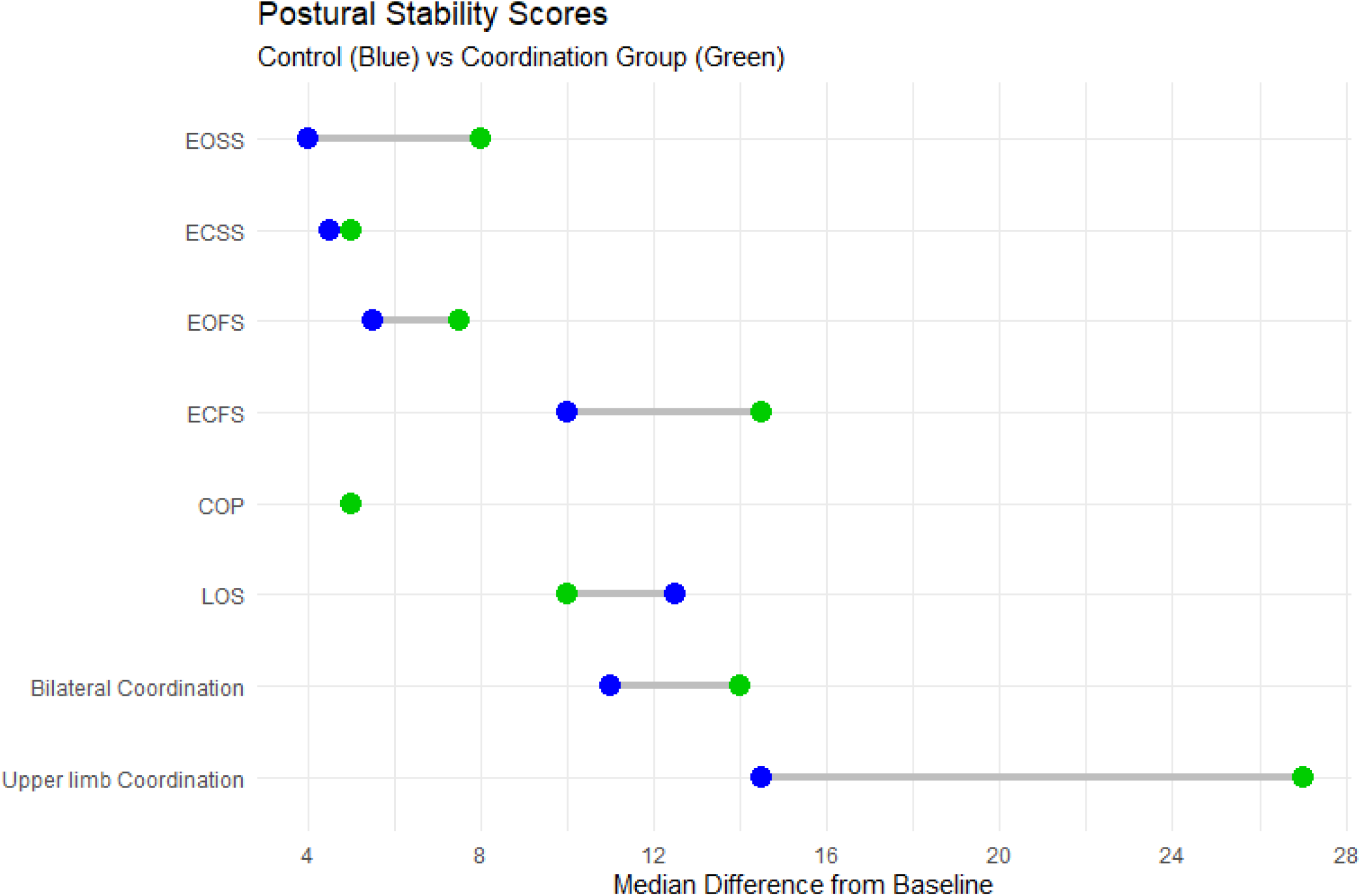
Postural stability and coordination scores

## 4. DISCUSSION

The current study was designed to investigate how physical exercise affected postural stability and coordination in children with posterior fossa tumors. during the maintenance phase of treatment, aiming to create a clear and precise understanding of their impairments. The present study included posterior fossa tumor that constitutes a major classification among brain tumor types. This was supported by **Packer (1999)**^19^ who stated that Medulloblastoma is the most common malignant brain tumor in pediatric patients and is a significant cause of cancer morbidity in children.

In the current study, choosing the age of children between 5 and 12 years old was in agreement with **Eccles (1999)**^20^ who reported that child starting to have a good performance like adult between the age of 5 and 12 years. More attention had to be paid to understand the potentially presenting side effects resulting from the administrated treatment or the disease itself, especially postural stability and coordination problems which were one of the least investigated problems among posterior fossa tumor patients.^21^

Participation of children with posterior fossa tumor in the rehabilitation program was associated with improved postural stability, coordination, and indirectly affect their overall physical performance of their daily living activities. The current study found improvements in the control group’s HUMAC balance and tilt system and BOT-2 scores before and after therapy; that was supported by **Mix et al. (2017)**^22^ who revealed evidence that completed rehabilitation programs increase mobility function in patients with brain tumors at a rate equivalent to motor function rehabilitation treatment in those with benign neurological disorders. Participation in a rehabilitation process is also linked to improved execution in day-to-day personal activities in patients with brain tumors.

Core stability exercises improve abdominal muscle workouts which have been suggested as a way to improve spine stability. Using the clinical Pilates approach improves abdominal muscle activation and increases intra-abdominal pressure, which leads to better functional outcomes and overall general activity.^23^

Regular exercise, in general, may be useful prior to, during, and after cancer treatment. Exercise improves the performance of body and mind, fatigue can be reduced, which can help with depression and anxiety. Exercise maintains or strengthen physical abilities to complete tasks. Muscle strength, bone health, and range of motion can all be improved. Exercise can help achieve and maintain a healthy weight by boosting your immune system and appetite. It also reduces treatment adverse effects and improve quality of life.^24^

The considerable improvement in the post treatment mean values of the measured variables of the postural stability group was attributed to the influence of postural stability exercises conducted on the HUMAC balance and tilt system. This may be due to the fact that proper balance demands multisystem feedback as well as healthy cerebellar and cognitive functions. The more challenging the work, the more cerebellar control and cognitive ability are assumed to be required.^9^ HUMAC balance and tilt system workouts are statistically proved to improve postural stability, according to the current study. Balance exercises are safe and useful for developing both static and dynamic balance, as well as enhancing motor skills, kinesthetic awareness, proprioception, and core muscles in youngsters, according to research. Finally, it improves the psychological well-being of cancer patients’ children.^25^

In order to beat cancer, a patient must not only beat the disease, but also deal with the therapy’s side effects.^26^ In the current study, physical activity appears to have beneficial impacts on numerous elements of the disease, its sequelae, and the therapy’s side effects, according to emerging research. For example, weaker muscles can be successfully strengthened, and there have been reports of good benefits on fatigue syndrome, a typical side effect of cancer treatment.^27^

The considerable improvement in the post-treatment mean values of the evaluated variables of the coordination group was attributable to the influence of the Bruininks-Oseretsky Test of Motor Proficiency coordination exercises. This might be due to the fact that coordination training provides a lot of health and well-being benefits for children. It helps them improve their technique and form, as well as their emotions and mental health, and reduces the risk of injury in the future. Coordination exercises can assist in the development of muscle, boost daily energy levels, increase flexibility and agility, improve memory and concentration, and stimulate the release of endorphins (called happy hormones).^28,29^

Somatosensory, visual, and vestibular information, as well as a completely coordinated neuromuscular system from the motor brain to the spinal cord, are all necessary for inducing a coordinated response. The muscles’ speed, distance, direction, timing, and precision are all features of coordinated action. They also have comparable synergistic effects (muscle recruitment), easy reversal between opposing muscle groups (the proper contraction and relaxation sequence), and proximal fixation, which can help with distal movement or posture maintenance.^30^

From the obtained results of the current study, it can be concluded that Postural stability and coordination improve physical activity in children with posterior fossa tumors.

## Data Availability

All data produced in the present work are contained in the manuscript

## DATA AVAILABILITY STATEMENT

The data that support the findings of this study are available from the corresponding author upon reasonable request.

