## Supplementary material for "IMPACT OF PHYSICAL ACTIVITY ON POSTURAL STABILITY AND COORDINATION IN CHILDREN WITH POSTERIOR FOSSA TUMOR: RANDOMIZED CONTROL PHASE III TRIAL": TABLE 1

**TABLE 1 Demographic and clinical characteristics**

| Items | | N (%) |
| --- | --- | --- |
| Age (Median, IQR) | | (7.78, 2.12) |
| Gender | Males | 38 (63%) |
|  | Females | 22 (37%) |
| Diagnosis | Low Grade Glioma | 29 (48.3%) |
|  | Medulloblastoma | 24 (40%) |
|  | Ependymoma | 3 (5%) |
|  | Brainstem Glioma | 2 (3.3%) |
|  | Other Diagnoses* | 2 (3.3%) |
| Treatment protocol | EPND (CCHE 3-1-2008) | 3 (5%) |
|  | Follow-up only | 30 (50%) |
|  | HRMB (CCHE 3-1-2008) | 11 (18.3%) |
|  | LGG (CCHE 3-1-2008) | 2 (3.3%) |
|  | RTH | 1 (1.7%) |
|  | SRMB (CCHE 3-1-2008) | 13 (21.7%) |
| Extent of resection | Gross/Near total resection | 51 (85%) |
|  | Subtotal resection | 3 (5%) |
|  | Biopsy | 3 (5%) |
|  | No surgery | 3 (5%) |
| Median time since end of treatment | | 17.5 months |

* One Non-biopsied SOL case and the other Choroid Plexus tumor.

**EPND**: Ependymoma, **HRMB**: high risk medulloblastoma **LGG**: Low grade glioma, **RTH**: Radiotherapy, **SRMB**: standard risk medulloblastoma.
