## Supplementary material for "IMPACT OF PHYSICAL ACTIVITY ON POSTURAL STABILITY AND COORDINATION IN CHILDREN WITH POSTERIOR FOSSA TUMOR: RANDOMIZED CONTROL PHASE III TRIAL": TABLE 2

**TABLE 2 Comparison between groups post intervention**

|  | Median ‘Pre-intervention’ | Median ‘Post-intervention’ | IQR (Tukey’s Hinges) | Adjusted P-value* |
| --- | --- | --- | --- | --- |
| **EOSS** |  |  |  |  |
| Control | 84 | 88 | 86 – 88.5 | — |
| Postural stability group | 79 | 90 | 89 – 90 | < 0.0001 |
| Coordination group | 77.5 | 85.5 | 85 – 89 | 0.019 |
| **ECSS** |  |  |  |  |
| Control | 77 | 80 | 79.5 – 85 | — |
| Postural stability group | 74 | 87.5 | 85 – 88 | 0.0002 |
| Coordination group | 75 | 79.5 | 76.5 – 86.5 | 0.92 |
| **EOFS** |  |  |  |  |
| Control | 58.5 | 68.5 | 79 – 90 | — |
| Postural stability group | 60.5 | 80 | 77 – 85 | < 0.0001 |
| Coordination group | 64.5 | 72.5 | 62.5 – 78 | 0.43 |
| **ECFS** |  |  |  |  |
| Control | 36.5 | 49 | 35 – 70 | — |
| Postural stability group | 39 | 71.5 | 67.5 – 75 | < 0.0001 |
| Coordination group | 46.5 | 60 | 50 – 68 | 0.16 |
| **COP** |  |  |  |  |
| Control | 80.5 | 85.5 | 85- 89.5 | — |
| Postural stability group | 83 | 93 | 89 – 95 | < 0.0001 |
| Coordination group | 84 | 90 | 88 – 90 | 0.15 |
| **LOS** |  |  |  |  |
| Control | 22.5 | 36 | 30 - 40 | — |
| Postural stability group | 25 | 51.5 | 44 – 54 | < 0.0001 |
| Coordination group | 26 | 40 | 30.5 – 44 | 0.56 |
| **Bilateral Coordination** |  |  |  |  |
| Control | 8 | 18 | 17.5 – 19 | — |
| Postural stability group | 7 | 20 | 19 – 20 | 0.0012 |
| Coordination group | 8 | 22 | 21 – 22 | < 0.0001 |
| **Upper Limb Coordination** |  |  |  |  |
| Control | 10 | 25 | 22 – 31.5 | — |
| Postural stability group | 9.5 | 30 | 29.5 – 32 | 0.005 |
| Coordination group | 10 | 36 | 35 – 37 | < 0.0001 |

* All p-values are adjusted for baseline measurements using a cumulative probability model.
